# The Gut-Heart Axis in Systemic Sclerosis: Evidence from the GENISOS cohort

**DOI:** 10.1101/2025.11.14.25340168

**Authors:** FR Di Ciommo, AP Balar, A Strother, S Kulkarni, M Hughes, B Skaug, MD Mayes, S Assassi, AY Ayla, ZH McMahan

**Author notes:** Statement of equal contribution: Ali Y. Ayla and Zsuzsanna McMahan contributed equally to this study. Corresponding author details: Zsuzsanna McMahan, MD, MHS, 6431 Fannin St., MSB 5.281, Houston, TX 77030.

## Abstract

**Background:** Cardiac involvement significantly impacts prognosis in systemic sclerosis (SSc), highlighting the need for early risk stratification. Gastrointestinal (GI) symptoms are common and often manifest early. Emerging data suggest a link between GI and cardiac manifestations, possibly through shared mechanisms like dysautonomia. This study investigates the overall association between GI and cardiac involvement in early SSc and evaluates whether baseline GI symptoms predict future cardiac manifestations.

**Methods:** We analyzed 459 patients from the prospective GENISOS cohort. GI involvement at baseline was defined by one or more of the following: dysphagia, peptic ulcer, bloating, diarrhea, malabsorption, constipation, or pseudo-obstruction. Cardiac manifestations included conduction defects and systolic dysfunction. Cox and multivariable logistic regression models assessed associations, adjusting for potential confounders.

**Results:** At baseline, 59% of patients had GI involvement. During follow-up, 26% of patients developed cardiac manifestations—mainly conduction defects (24%) and less commonly systolic dysfunction (5%). Baseline malabsorption and bloating were strong predictors of future cardiac involvement, with malabsorption showing the highest risk [HR 12.38 (95% CI: 4.4 – 34.6)]. Interestingly, dysphagia and peptic ulcers were significantly associated with conduction defects, while malabsorption was significantly associated with systolic dysfunction, even after adjustment for potential confounders.

**Conclusions:** Upper GI dysfunction was associated specifically with conduction defects, suggesting that autonomic dysfunction contributes. In contrast, lower GI involvement, particularly malabsorption, was linked to systolic dysfunction in SSc patients, potentially indicating a distinct biological mechanism. These findings may support integrating early GI symptoms into cardiac risk stratification, and provide a foundation for future translational studies.

**Key messages:**

- Distinct cardiac phenotypes in SSc patients associate with distinct types of GI involvement.
- Upper GI manifestations were significantly associated with conduction defects, possibly reflecting shared dysautonomia, affecting both GI motility and cardiac rhythm.
- Lower GI involvement was significantly associated with systolic dysfunction, suggesting that a distinct mechanism may underlie this clinical phenotype.

## INTRODUCTION

Systemic sclerosis (SSc) is a chronic autoimmune disease characterized by progressive vasculopathy and fibrosis. With progression, multiorgan involvement leads to heterogeneous clinical manifestations, significant disability, and often mortality(1, 2). Among its complications, the gastrointestinal (GI) tract is one of the most commonly affected, impacting nearly 90% of patients. SSc can impair the entire digestive tract, from the oropharynx to the anorectum, resulting in a wide spectrum of symptoms (3–5). Despite its high prevalence, the underlying pathogenic mechanisms remain largely unknown, posing challenges for clinical monitoring and management.

A growing body of evidence suggests a link between GI and cardiac complications in SSc. Notably, dysautonomia, which is believed to contribute to upper GI dysfunction and cardiac involvement (specifically dysrhythmias and conduction disturbances), is highly prevalent in patients with SSc-GI disease (6, 7). Furthermore, data from the EUSTAR cohort found that general intestinal symptoms were associated with an increased risk of new SSc-primary heart involvement (pHI) (8). Finally, emerging research also demonstrates that the mesoderm-derived enteric neurons (MENS) are specifically targeted by the scleroderma autoimmune response, which may also impact mesoderm-derived tissue in cardiac muscle (9, 10). This has raised questions about a deeper biological connection or shared susceptibility between mesoderm-derived enteric neurons and other mesodermal-derived tissues (11).

Given the prognostic significance of cardiac involvement in SSc, we aimed to determine whether specific baseline GI manifestations could serve as early indicators of distinct types of future cardiac involvement. While GI involvement is common, its predictive value for specific cardiac outcomes has not been thoroughly explored. Here, we utilized a large, well-characterized cohort of patients with early SSc to: (1) clarify the temporal association between GI and cardiac involvement; and (2) define the clinical phenotype(s) of these high-risk patient subsets in early disease.

## PATIENTS AND METHODS

### Patient Cohort

The Genetics versus Environment in Scleroderma Outcomes Study (GENISOS) is a multicenter, prospective cohort study involving UTHealth Houston, UTHealth San Antonio, and UTMB Galveston, focusing on well-characterized early SSc patients. Recruitment began in January 1998 and is ongoing, with a maximum follow-up of 20 years. As previously described (12, 13), patients were eligible if they met the following criteria: age >18 years at enrollment, fulfillment of either the 2013 ACR/EULAR classification criteria (14) or the 1980 ACR criteria (15), disease onset (first non-Raynaud symptom) within five years prior to enrollment. Patient-reported demographic, psychological, and clinical data were collected at baseline and at scheduled follow-up visits, every six months for the first two years, followed by annual assessments.

All participants signed written informed consent, and ethical approval was obtained from the UTHealth Houston Institutional Review Boards.

### Clinical phenotyping of SSc

Clinical data included age, sex, race, ethnicity, and smoking status. Disease duration was defined as the interval between the first non-Raynaud’s symptom and the last documented follow-up. Cutaneous subtype was classified as diffuse or limited based on the extent of skin thickening (16). The presence of telangiectasia and pulmonary involvement was documented over time. Restrictive lung disease was assessed using pulmonary function tests (PFTs), including forced vital capacity (FVC) and single-breath diffusing capacity for carbon monoxide (DLCO), recorded as absolute and percentage-predicted values standardized for sex, age, and race. Analyses at baseline and over time were performed using the minimum predicted values for these parameters. Pulmonary hypertension was documented by echocardiography or right heart catheterization (mean PA pressure ≥ 25 mmHg or systolic PAP ≥ 40 mmHg). Sicca symptoms were defined as the presence of at least one of the following: dry eyes and/or mouth lasting more than three months, sensation of sand in the eyes, use of artificial tears at least three times daily, salivary glands swelling, or needing liquids to swallow due to dry mouth.

### GI involvement

GI manifestations were documented longitudinally by the treating physician and consisted of one or more of the following findings: (a) dysphagia (defined as pain or difficulty of swallowing or passing food); (b) clinically significant peptic ulcers (documented per esophagogastroduodenoscopy); (c) malabsorption (confirmed by diarrhea with > 10% weight loss or by laboratory evidence of nutritional deficiencies); (d) pseudo-obstruction (diagnosed through radiographic studies); (e) bloating; (f) diarrhea; and (g) constipation, per patient report. In this manuscript, we use the term “GI involvement” to define the presence of at least one of the manifestations listed above. The term “GI manifestation” refers to one specific GI symptom.

### Cardiac involvement

To focus on objectively measured data obtained through diagnostic tools, we defined “cardiac involvement” as the occurrence of at least one of the following two manifestations: (a) systolic dysfunction, documented by echocardiography (ejection fraction < 40%) or nuclear imaging; (b) conduction defects, identified via resting or ambulatory electrocardiography. These variables are captured longitudinally in our Center’s database. In this manuscript, the term “cardiac manifestation” refers to either of the two conditions defined above.

To capture clinical phenotype, all analyses were performed using the maximum recorded value for continuous variables. Dichotomous variables were categorized as “ever/never,” meaning that “ever” is equivalent to a patient having a given condition (e.g., systolic dysfunction) at any time during the follow-up period.

### Autoantibody profiles

Baseline serum samples were analyzed at UTHealth Houston. Anti-nuclear antibodies (ANA) and anti-centromere antibodies (ACA) were detected using indirect immunofluorescence with Hep-2 cell substrates (Antibodies Incorporated, Davis, CA, USA). Anti-topoisomerase I (ATA) and anti-ribonucleoprotein antibodies were identified using passive immunodiffusion against calf thymus extract (INOVA Diagnostics, San Diego, CA, USA). Anti-RNA polymerase III antibodies were measured using ELISA (MBL, Nagoya, Japan).

### Statistical methods

Data distribution was initially assessed using box plots and histograms. A dichotomous variable, “Any GI”, was defined by the presence (yes/no) of at least one gastrointestinal manifestation during the disease course: dysphagia, peptic ulcer, bloating, diarrhea, constipation, malabsorption, or pseudo-obstruction. Similarly, “Any Heart” indicated the presence of any previously defined cardiac manifestation (systolic dysfunction and/or conduction defects) during follow-up.

A baseline descriptive analysis was followed by a longitudinal comparison of patients with and without GI involvement. Patients could present with multiple conditions within the Any GI and/or Any Heart categories, each recorded and analyzed independently. For the analyses, only patients with complete data across both the GI and cardiac domains were included; patients with missing data were excluded. Continuous variables were compared using Student’s t-test (parametric) or Wilcoxon signed-rank test (non-parametric), while associations between dichotomous variables were assessed using Fisher’s exact or Chi-square tests. Results are reported as n (%) for categorical variables, mean (SD) for normally distributed continuous variables, and median (IQR) for non-normally distributed ones.

Kaplan–Meier curves and log-rank tests were used to evaluate the association between baseline GI involvement and cardiac manifestations. Patients with cardiac manifestations that occurred at baseline and those with cardiac events occurring before or concurrently with GI involvement were excluded from our time-to-event analysis. To explore temporal relationships, Cox proportional hazards models were applied, reporting hazard ratios (HRs), 95% confidence intervals (CIs), and p-values (significance set at p < 0.05). Univariate HRs were first calculated, then adjusted for significant variables and established confounders.

Univariable regression was used to assess associations, followed by a multivariable model incorporating significant univariate results (p < 0.05) and known confounders from the literature (17–21). In the adjusted analysis, we calculated age and disease duration at the time of the first cardiovascular event or, for those without events, the maximum age and disease duration reached during the study period. All analyses were conducted using Stata version 18.0.

## RESULTS

### Baseline characteristics of the cohort

Our cohort consisted of 459 SSc patients, 378 (82%) females, with a median follow-up time of 4.10 years (IQR = 0.8 – 8.1) and a median number of recorded visits of 5 (IQR = 1 – 9). At baseline, the mean age was 48 ± 13 years, and the median disease duration from the first non-Raynaud symptom was 2.4 years (IQR = 1.3 – 3.6). The diffuse cutaneous subtype was the most common, accounting for 61% of cases. GI involvement was present at baseline in 273 out of 459 patients (59%). The median time from the first recorded visit to the onset of cardiac complications was 1.12 years (IQR: 0 – 4.96), with some cases occurring as late as 14.40 years into follow-up. Among the studied cohort, 83 out of 447 patients (19%) tested positive for anti-topoisomerase I antibodies, 59 out of 448 (13%) for anti-centromere antibodies, 98 out of 448 (22%) for anti-RNA polymerase III antibodies, 36 out of 439 (8%) for anti-RNP antibodies, and 25 out of 102 (24%) for anti-U3RNP antibodies. All results are presented in Table 1.

**Table 1.**
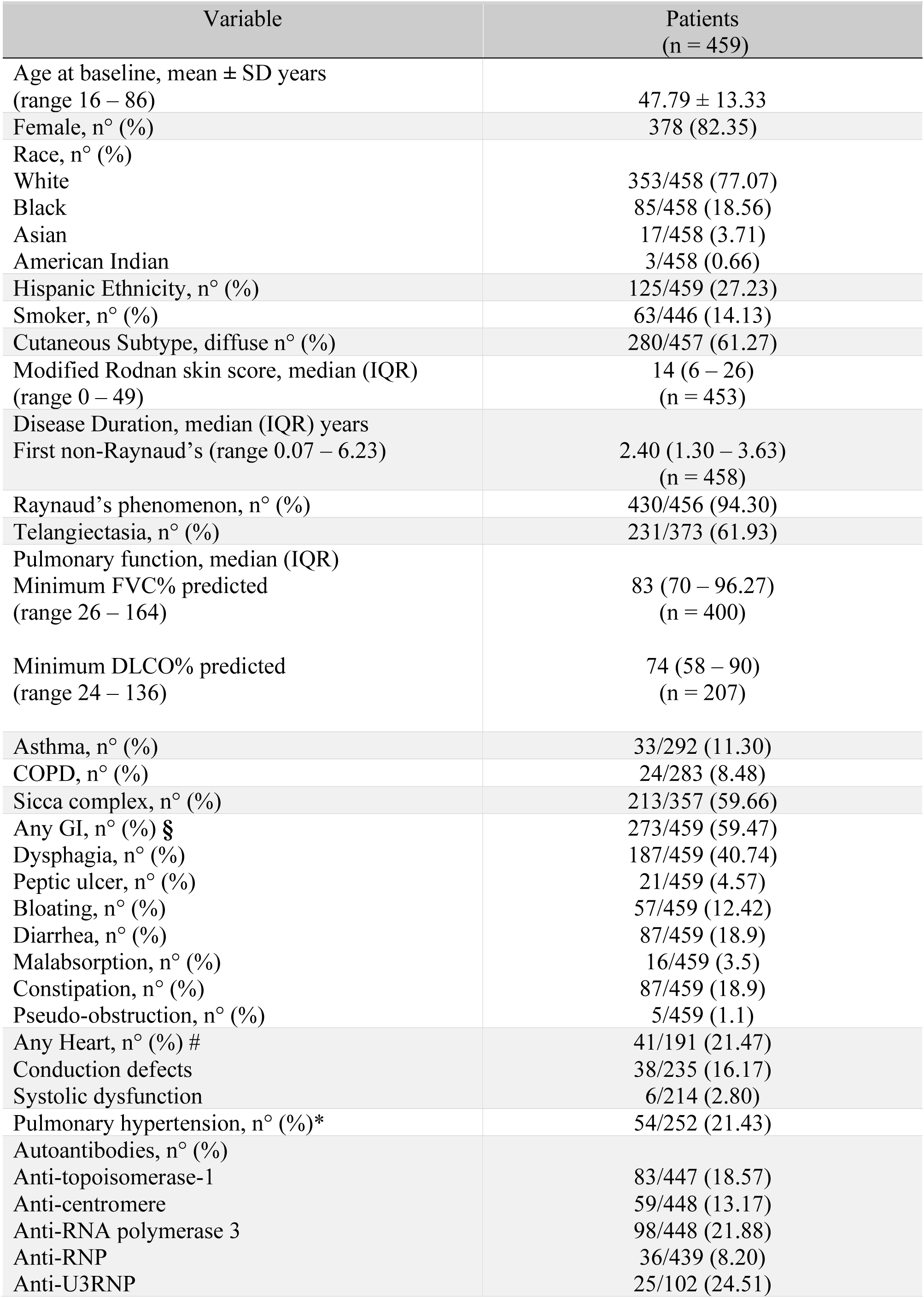
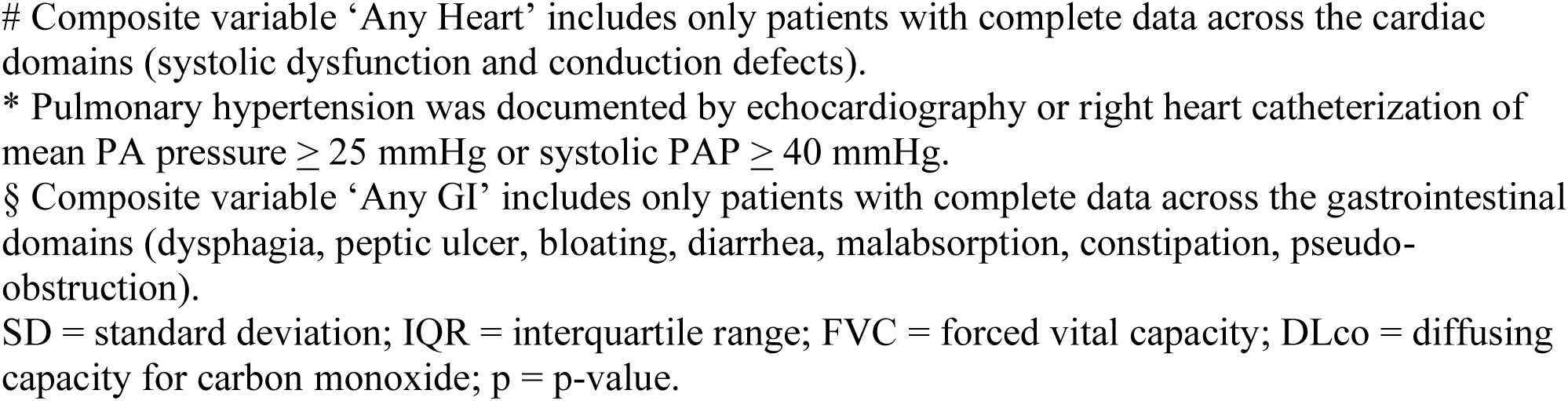
Demographic, clinical, and autoantibody characteristics of SSc patients from the GENISOS cohort at baseline.

### Early symptoms of bloating and malabsorption are significant predictors of subsequent cardiac complications in SSc

In our cohort, 106 out of 459 patients (23%) developed both cardiac (‘any heart’) and GI (‘any GI) manifestations during the course of SSc. In 64 of these cases (60%), GI manifestations preceded the onset of cardiac complications, while in 10 cases (9%), cardiac complications occurred first. For the remaining 32 patients (30%), both manifestations emerged simultaneously.

Given that GI involvement frequently precedes the onset of cardiac manifestations, we initially sought to determine whether, and which, baseline GI manifestations might serve as early predictors of subsequent cardiac involvement. Interestingly, baseline GI involvement at the time of enrollment in the early SSc cohort was significantly associated with an increased risk of developing cardiac manifestations (log-rank p = 0.039). In particular, bloating and malabsorption emerged as strong predictors of future cardiac involvement, each showing a robust association with increased risk (log-rank p < 0.001). Other baseline GI manifestations, such as dysphagia, peptic ulcer, and diarrhea, were not predictive of future cardiac events (log-rank p >0.05). In the Cox regression analysis, baseline malabsorption was specifically associated with the highest risk of future cardiac events (HR = 19.90; 95% CI: 9.9 – 40.1). After adjusting for age, sex, race, and disease duration, both bloating and malabsorption at baseline remained significant predictors of increased risk of future cardiac manifestations (HR = 3.6, 95% CI: 2.2 – 5.7, and HR = 12.4, 95% CI: 4.4 – 34.6, respectively; see Table 2).

**Table 2.** Baseline GI predictors of cardiac risk *in patients with systemic sclerosis.

| Characteristic | Cox regression | Cox regression adjusted for age§, sex, race and disease duration# |
| --- | --- | --- |
| Any GI | **1.45 (1.02 – 2.07) †** | **1.48 (1.03 – 2.14) †** |
| Upper GI | 1.19 (0.84 – 1.67) | 1.02 (0.71 – 1.45) |
| *Dysphagia* | 1.24 (0.88 – 1.75) | 1.10 (0.77 – 1.56) |
| Lower GI | **1.82 (1.30 – 2.56) †** | **2.09 (1.45 – 2.99) †** |
| *Bloating* | **2.79 (1.91 – 4.10) †** | **3.57 (2.25 – 5.67) †** |
| *Diarrhea* | 1.06 (0.69 – 1.64) | 0.81 (0.49 – 1.34) |
| *Malabsorption* | **19.90 (9.89 – 40.06) †** | **12.38 (4.42 – 34.63) †** |

Definition of GI predictors: (a) Dysphagia, defined as pain or difficulty in swallowing or passing food; (b) Bloating; (c) Diarrhea; (d) Malabsorption, confirmed by diarrhea associated with >10% weight loss or by abnormal chemical studies. Symptoms (b) and (c) were self-reported by patients.
\* Cardiac risk refers to the risk of systolic dysfunction and/or conduction defects.
Values are Hazard Ratios (95% confidence interval).
§ Refers to the age at baseline.

### GI involvement and cardiac manifestations are bi-directionally linked across the SSc cohort

Though we initially determined that specific GI manifestations were baseline predictors of SSc cardiac involvement, we hypothesized that both cardiac and GI dysfunction in SSc are linked bidirectionally through SSc autoimmune responses to common tissues in both organs (i.e., neural crest and/or mesodermal-derived tissues), rather than by causality (i.e., GI involvement causing cardiac involvement) (6, 7, 9, 10). We therefore utilized data from our well-characterized cohort of patients with early SSc to determine whether individuals who had GI involvement at any time point were more likely also to experience SSc-related cardiac dysfunction during their disease course. Clinical, demographic, and autoantibody characteristics of the cohort, stratified by GI involvement, are summarized in Table 3. Importantly, over the course of follow-up, among the 459 patients, 118 (26 %) experienced at least one cardiac manifestation (any heart’), including 22 patients with systolic dysfunction (5%), 111 with conduction defects (24%), and 15 (3%) of patients presenting with both.

**Table 3.**
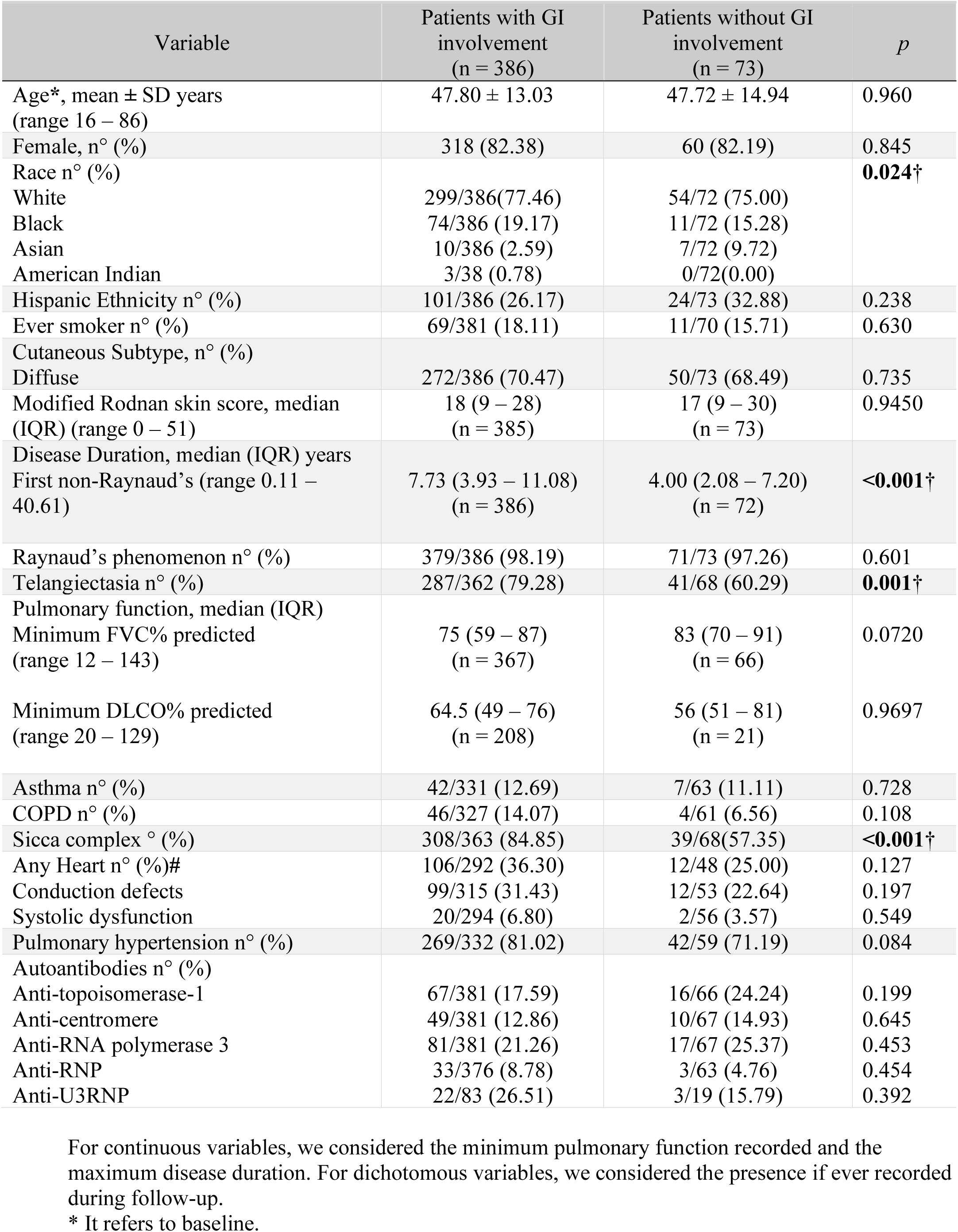
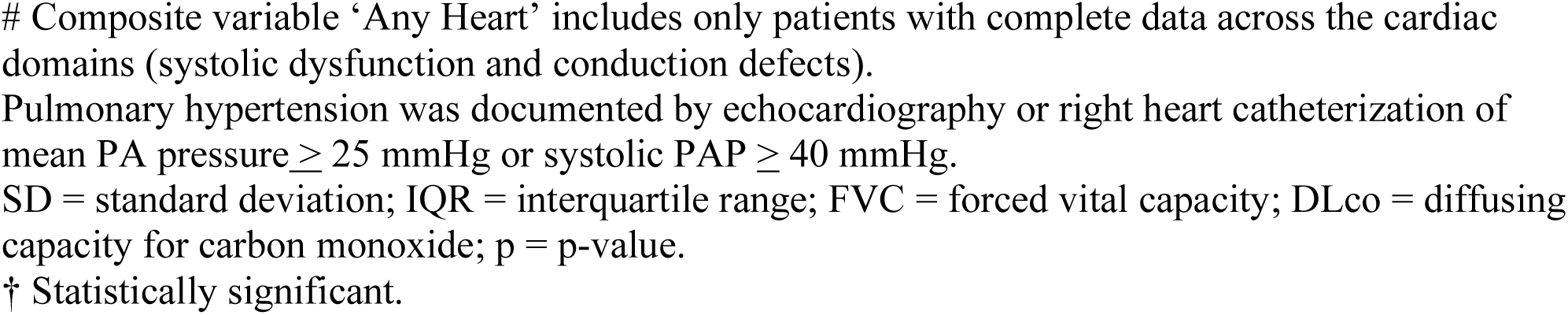
Demographic, clinical, and autoantibody profiles of SSc patients with and without GI involvement over time.

While the overall prevalence of cardiac involvement was numerically higher among those with GI disease (36% vs. 25%), this difference did not reach statistical significance (p = 0.127). Nonetheless, both conduction defects and systolic dysfunction occurred more frequently in the GI subgroup (31% vs. 23%, p = 0.197; and 7% vs. 4%, p = 0.549, respectively) (see Table 3).

Although the bivariate analysis did not reach statistical significance, the data revealed an interesting trend, with cardiac involvement being more frequent among patients with GI manifestations. Acknowledging both this pattern and the potential limitations of considering heterogenous GI involvement as a single group, we further explored the association between cardiac involvement and each specific GI manifestation.

Interestingly, we identified a significant association between specific GI manifestations and cardiac involvement. Patients with cardiac involvement had significantly higher rates of GI involvement, including dysphagia (76% vs. 61%; p = 0.005), peptic ulcers (19% vs. 7%; p = 0.001), and bloating (62% vs. 40%, p <0.001), as well as diarrhea (55% vs. 44%; p = 0.045) and malabsorption (17% vs. 7%, p = 0.005) than those without (see Table 4).

**Table 4.** Demographic, clinical, and autoantibody profiles of SSc patients with and without cardiac involvement over time.

| Variable | Patients with cardiac involvement<br>(n = 118) | Patients without cardiac involvement<br>(n = 222) | p |
| --- | --- | --- | --- |
| Age*, mean ± SD year<br>(range 16 – 86) | 50 ± 11.29 | 46.79 ± 13.33 | **0.030**† |
| Female, n° (%) | 87 (74.36) | 190 (85.59) | **0.011**† |
| Race, n° (%) |  |  | **0.003**† |
| White | 77/117(65.81) | 179/222 (80.63) |  |
| Black | 35/117 (29.91) | 32/222 (14.41) |  |
| Asian | 4/117 (3.42) | 11/222 (4.95) |  |
| American Indian | 1/117 (0.85) | 0/222(0.00) |  |
| Hispanic Ethnicity, n° (%) | 23/118 (19.49) | 56/222 (25.23) | 0.233 |
| Ever smoker, n° (%) | 27 /118 (22.88) | 36/219 (16.44) | 0.148 |
| Cutaneous Subtype, n° (%) |  |  |  |
| Diffuse | 89/118 (75.42) | 151/222(68.02) | 0.154 |
| Modified Rodnan skin score, median<br>(IQR) (range 0 – 51) | 20.5 (12 – 31)<br>(n = 118) | 16 (9 – 28)<br>(n = 222) | **0.018**† |
| Disease Duration, median (IQR) years<br>First non-Raynaud’s (range 0.11 –<br>40.61) | 8.34 (5.15 – 13.54)<br>(n = 118) | 8.56 (4.75 – 11.56)<br>(n = 222) | 0.7055 |
| Raynaud’s phenomenon, n° (%) | 117/118 (99.15) | 218/222 (98.20) | 0.662 |
| Telangiectasia, n° (%) | 96/116 (82.76) | 160/220 (72.73) | **0.040**† |
| Pulmonary function, median (IQR)<br>Minimum FVC% predicted<br>(range 12 – 143) | 72.1(57 – 84)<br>(n = 115) | 78 (62 – 91)<br>(n = 215) | **0.002**† |
| Minimum DLCO% predicted<br>(range 20 – 129) | 63 (46 – 70)<br>(n = 73) | 66.5 (50 – 79)<br>(n = 94) | 0.137 |
| Asthma, n° (%) | 15/111 (13.51) | 23/220 (10.45) | 0.410 |
| COPD, n° (%) | 21/111 (18.92) | 23/219 (10.50) | **0.034**† |
| Sicca complex, n° (%) | 103/116 (88.79) | 168/221(76.02) | **0.005**† |
| Any GI, n° (%)# | 106/118 (89.83) | 186/222 (83.78) | 0.127 |
| Dysphagia | 90/118 (76.27) | 136/222 (61.26) | **0.005**† |
| Peptic ulcer | 22/118 (18.64) | 16/222 (7.21) | **0.001**† |
| Bloating | 73/118 (61.86) | 89/222 (40.09) | **<0.001**† |
| Diarrhea | 65/118 (55.08) | 97/222 (43.69) | **0.045†** |
| Malabsorption | 20/118 (16.95) | 16/222 (7.21) | **0.005†** |
| Constipation | 58/118 (49.15) | 93/222 (41.89) | 0.200 |
| Pseudo-obstruction | 9/118 (7.63) | 9/222 (4.05) | 0.161 |
| Autoantibodies, n° (%) |  |  |  |
| Anti-topoisomerase-1 | 26/115 (22.61) | 37/218 (16.97) | 0.212 |
| Anti-centromere | 9/115 (7.83) | 34/218 (15.60) | **0.044†** |
| Anti-RNA polymerase 3 | 27/115 (23.48) | 41/218 (18.81) | 0.315 |
| Anti-RNP | 7/115(6.09) | 20/215 (9.30) | 0.310 |
| Anti-U3RNP | 13/38 (34.21) | 6/51 (11.76) | **0.017†** |

Cardiac involvement refers to the following domains: systolic dysfunction and conduction defects. For continuous variables, we considered the minimum pulmonary function recorded and the maximum disease duration. For dichotomous variables, we considered the presence if ever recorded during follow-up.
\* It refers to baseline.

We then sought to determine whether specific cardiac manifestation(s) were driving these associations. Interestingly, we found that patients with conduction defects had significantly more frequent upper GI symptoms, including dysphagia (76% vs. 61%, p = 0.007) and peptic ulcers (20% vs. 7%, p = 0.001). In addition, significant associations were observed with diarrhea (55% vs. 42%, p = 0.027) and malabsorption (17% vs. 7%, p = 0.003). In contrast, patients with systolic dysfunction had a higher prevalence of lower GI symptoms, including bloating (68% vs. 45%, p = 0.033) and malabsorption (32% vs. 8%, p <0.001) (data not shown).

### Quantifying the strength of the associations using univariate and multivariable logistic regression

Given that the bivariate analysis revealed that cardiac manifestations are more frequently encountered among SSc patients with GI involvement, we then sought to examine the strength of these associations. We therefore applied an unadjusted logistic regression model (Model 1), and then a multivariable model adjusting each GI manifestation separately for age, sex, race, and disease duration (Model 2).

In the unadjusted model (Model 1), strong associations were again identified between several GI manifestations and cardiac involvement, with peptic ulcer showing the strongest association (see Table 5). Specifically, patients with peptic ulcers had nearly three times the odds of exhibiting cardiac involvement compared to patients without peptic ulcers (OR: 2.9; 95% CI: 1.5 – 5.9).

**Table 5.** Associations between GI manifestations and cardiac involvement * in SSc patients over time: results from logistic models.

| Characteristic | Univariate logistic Regression: Model 1 | Conditional logistic adjusted for age §, sex, race, and disease duration#: Model 2 |
| --- | --- | --- |
| Age, (years) § | 0.99 (0.98 – 1.02) | 1.02 (0.99 – 1.04) |
| Sex, (female) | **0.49 (0.28 – 0.85) †** | **0.32 (0.14 – 0.75) †** |
| Race, (black) | **2.54 (1.47 – 4.40) †** | **3.84 (1.54 – 9.58) †** |
| Disease duration# | **0.78 (0.72 – 0.84) †** | **0.72 (0.66 – 0.80) †** |
| Modified Rodnan skin score | **1.02 (1.00 – 1.04) †** | 1.02 (0.99 – 1.05) |
| Telangiectasia | **1.8 (1.02 – 3.17) †** | 1.80 (0.75 – 4.32) |
| Minimum FVC % predicted | **0.98 (0.97 – 0.99) †** | 0.99 (0.97 – 1.00) |
| COPD | **1.99 (1.05 – 3.78) †** | 1.46 (0.59 – 3.63) |
| Sicca | **2.50 (1.30 – 4.81) †** | **5.41 (1.95 – 15.00) †** |
| Dysphagia | **2.03 (1.23 – 3.36) †** | 1.73 (0.80 – 3.73) |
| Peptic ulcer | **2.95 (1.48 – 5.87) †** | **3.74 (1.36 – 10.30) †** |
| Bloating | **2.42 (1.53 – 3.83) †** | **3.10 (1.54 – 6.27) †** |
| Diarrhea | **1.58 (1.01 – 2.48) †** | 0.82 (0.39 – 1.74) |
| Malabsorption | **2.63 (1.30 – 5.29) †** | 1.48 (0.46 – 4.79) |
| Anti-U3RNP | **3.9 (1.32 – 11.53) †** | 1.04 (0.70 – 1.55) |

Definition of GI manifestations: (a) Dysphagia, defined as pain or difficulty in swallowing or passing food; (b) Peptic ulcer, documented; (c) Bloating; (d) Diarrhea; (e) Malabsorption, confirmed by diarrhea associated with >10% weight loss or by abnormal chemical studies.
Symptoms from (c) to (d) were self-reported by patients.
FVC = forced vital capacity.
\* Cardiac involvement refers to the following domains: systolic dysfunction and conduction defects.
§ Refers to the age at the time of the first cardiac event (if applicable) or the maximum age reached during the study period (if no cardiac event occurred).

Additionally, notable associations were observed for dysphagia (OR: 2.0; 95% CI:1.2 – 3.4), bloating (OR: 2.4; 95% CI: 1.5 – 3.8), diarrhea (OR: 1.6; 95% CI: 1.0 – 2.5), and malabsorption (OR: 2.6; 95% CI: 1.3 – 5.3). When adjusting for potential confounders in Model 2, only the associations of peptic ulcers (OR: 3.7; 95% CI: 1.4 – 10.3) and bloating (OR: 3.1; 95% CI: 1.5 – 6.3) with cardiac involvement remained strong, whereas the others lost statistical significance (see Table 5).

We next quantified the strength of associations specifically between (1) conduction defects, and (2) systolic dysfunction, and individual GI manifestations. In the unadjusted models, cardiac conduction defects were significantly more likely to occur in patients with dysphagia (OR: 1.9; 95% CI: 1.2 – 3.3), peptic ulcer (OR: 3.1; 95% CI: 1.6 – 5.9), bloating (OR: 2.2; 95% CI: 1.4 – 3.5), diarrhea (OR: 1.7; 95% CI: 1.1 – 2.6), and/or malabsorption (OR: 2.7; 95% CI: 1.4 – 5.5). In contrast, systolic dysfunction was only enriched among patients with bloating (OR = 2.6; 95% CI: 1.1 – 6.6) and/or malabsorption (OR = 5.2; 95% CI: 1.9 – 13.9). After adjustment in Model 2, peptic ulcer (OR: 4.4; 95% CI: 1.8–10.7) and bloating (OR: 2.3; 95% CI: 1.2–4.4) continued to show robust associations with cardiac conduction defects. In contrast, systolic dysfunction remained significantly associated with malabsorption (OR = 4.3; 95% CI: 1.4–13.4) in the adjusted model (data not shown).

## DISCUSSION

This study offers novel insights into the interplay between GI and cardiac involvement in early SSc, a relationship that has been largely overlooked despite the high prevalence of both complications. We demonstrate that distinct cardiac phenotypes are associated with specific patterns of GI involvement, suggesting divergent underlying mechanisms. Upper GI symptoms, such as dysphagia and peptic ulcer disease, were significantly associated with conduction abnormalities, pointing to a possible shared autonomic dysfunction affecting both GI motility and cardiac rhythm. In contrast, lower GI manifestations, particularly malabsorption, were linked to systolic dysfunction, potentially indicating a separate pathophysiologic pathway. To our knowledge, this is the first study to systematically characterize these associations, highlighting the need for integrated organ system assessment in early SSc and opening new avenues for risk stratification and targeted intervention.

The strong link between upper GI manifestations and conduction defects may reflect autonomic dysfunction, a well-known contributor to both upper GI dysmotility and cardiac arrhythmias, which is known to exist in SSc patients. Autonomic imbalance—characterized by increased sympathetic tone and reduced vagal activity—impairs upper GI motility and lower esophageal sphincter function, leading to esophageal and gastric dysmotility, dysphagia, and reflux (7). Our recent study found that SSc patients with global autonomic dysfunction (defined as >5 positive COMPASS-31 subdomains) had abnormal GI transit and higher COMPASS-31 scores, particularly in patients with upper GI involvement (6). Conduction defects, a hallmark of cardiac involvement in SSc, may also be caused or exacerbated by this autonomic imbalance (7, 22).

The strong association between peptic ulcer disease and conduction defects is particularly notable, given the vagus nerve’s role in controlling both autonomic regulation and gastric acid secretion. Vagal dysfunction may contribute to ulcer development and, through vagally mediated reflexes, promote rhythm disturbances. This is nicely demonstrated in patients with gastrocardiac (Roemheld) syndrome, where esophageal acid stimulation disrupts sympatho-vagal balance and induces arrhythmias (22, 23). Thus, upper GI dysfunction and conduction defects may represent parallel outcomes of shared autonomic impairment in SSc.

In contrast, the robust association between malabsorption and reduced ejection fraction heart failure (<40%) suggests a different pathophysiological mechanism. In our cohort, malabsorption was defined by persistent diarrhea, >10% unintentional weight loss, and laboratory evidence of nutritional deficiencies—capturing a chronic, clinically significant intestinal dysfunction. Three hypotheses may explain this association. First, emerging evidence suggests a link between gut dysbiosis and adverse cardiovascular outcomes (24, 25). The EUSTAR cohort recently reported an increased risk of primary heart involvement in SSc patients with intestinal symptoms such as bloating and diarrhea (8). Dysbiosis may impair intestinal barrier integrity by reducing short-chain fatty acid (SCFA) production, thereby allowing the translocation of pro-inflammatory bacterial products such as lipopolysaccharide (LPS) into the circulation. Elevated LPS levels have been observed in heart failure and correlate with congestion (24). Second, elevated levels of microbial metabolite trimethylamine N-oxide (TMAO)—derived from dietary choline and L-carnitine—are associated with adverse ventricular remodeling, impaired cardiac function, and increased mortality in heart failure (25). In SSc, elevated TMAO levels inversely correlate with left ventricular ejection fraction, suggesting a role in reduced systolic performance (26). These findings support the hypothesis that gut-derived metabolites may contribute to cardiac dysfunction in SSc. Finally, recent findings from our group have identified mesoderm-derived enteric neurons as targets of the autoimmune response in SSc. Given that the cardiac myocardium also originates from mesodermal tissue, this may point to a shared developmental vulnerability that contributes to parallel involvement of the enteric nervous system and cardiac muscle in SSc. These observations provide a compelling foundation for future translational studies to elucidate common pathogenic mechanisms and identify novel therapeutic targets across organ systems.

In our cohort, many patients reported GI involvement prior to cardiac complications. When we examined whether baseline GI manifestations predicted future cardiac involvement, bloating—and especially malabsorption—were significantly associated with subsequent cardiac involvement. In contrast, manifestations of upper GI disease, like dysphagia and peptic ulcers, while strongly associated with conduction defects, did not predict future cardiac complications. This pattern suggests distinct temporal and mechanistic relationships: upper GI symptoms and conduction defects may occur concurrently due to autonomic dysfunction, whereas malabsorption and systolic dysfunction may follow a sequential process. For example, chronic intestinal dysfunction may drive systemic inflammation and microbial translocation, contributing to myocardial damage. These findings underscore the importance of early cardiovascular risk stratification and close monitoring in SSc.

A major strength of our study is the large, well-characterized cohort of nearly 500 early SSc patients, followed for up to 20 years with comprehensive clinical and biological data. This robust longitudinal dataset enabled detailed evaluation of the temporal dynamics between GI and cardiac involvement and the potential bidirectionality of this relationship. To our knowledge, this is the first study to explore the relationship between distinct cardiac and GI manifestations in SSc with such depth, moving beyond the traditional focus on isolated organ systems to propose a more integrated disease model. Our findings not only support earlier cardiovascular risk stratification but also provide a framework for exploring shared pathophysiological mechanisms across organ systems. Importantly, they highlight the heterogeneous nature of the GI tract, suggesting that different segments may be affected independently through distinct mechanisms, each contributing uniquely to disease progression. This may have critical implications for both understanding disease pathogenesis and tailoring clinical management.

Several limitations should be noted. As detailed in the “Methods”, missing data for certain cardiac and GI variables may have reduced statistical power. Patient-reported outcomes were unavailable, as these measures were developed after most data collection for this older cohort. While cardiac involvement was objectively assessed using ECG and echocardiography, some GI manifestations were based on physician reports and may be underrepresented. Finally, although our results demonstrate a strong association between GI and cardiac involvement, functional and mechanistic studies are needed to confirm a causal or biologically mediated link between these systems.

In summary, this is the first study to provide evidence of a strong complex association between GI and cardiac involvement in SSc. Our findings move beyond a unidirectional risk model, revealing potential bidirectional interactions and supporting a more integrated, multiorgan perspective. By identifying distinct cardiac phenotypes associated with specific GI manifestations, our study underscores the heterogeneity of organ involvement in SSc and provides a framework for improving risk stratification, disease monitoring, and therapeutic approaches.

## Data Availability

All data produced in the present study are available upon reasonable request to the authors

